# Perceptions of precision health research participation: a cognitive interview study

**DOI:** 10.64898/2026.03.30.26349735

**Authors:** Rachael J. Werner, Samiha T. Karim, Melissa A. Cunningham, Lee H. Moultrie, Marquetta L. Goodwine, Lori Ann Ueberroth, Bethany J. Wolf, Caitlin G. Allen, Diane L. Kamen, Paula S. Ramos

## Abstract

**Background:** The Precision rEsearCh pArticipatioN (PECAN) study aims to explore factors that influence perceptions of precision health research participation - defined as research integrating genetic, behavioral, and environmental determinants to tailor individualized disease prevention and treatment - focusing on diverse communities in South Carolina. The objective is to identify factors influencing participation in precision health research and inform strategies to reduce barriers among populations disproportionately affected by health disparities.

**Methods:** To ensure the cultural accessibility and comprehensibility of the survey instrument for the PECAN study, researchers conducted a qualitative cognitive interviewing study using a purposive sampling strategy. Four African American community members from the Sea Island Families Project Citizen Advisory Committee participated in a 90-minute cognitive interview conducted using concurrent verbal probing. Participants reviewed a draft PECAN survey designed to assess perceptions of precision health research participation and provided feedback regarding question clarity, comprehension, cultural relevance, and ease of response. Four investigators independently recorded notes, identified recurring themes through consensus review, and used participant feedback to refine survey wording, structure, and content.

**Results:** The cognitive interview study identified several survey items that participants found challenging or ambiguous, particularly due to complex wording, culturally irrelevant content, and questions requiring extensive recall. Participants emphasized the need for clearer language, reassurance about anonymity, and the use of biological terms, as well as greater cultural representation. Based on participant feedback, revisions included simplifying language, clarifying terminology, adding contextual statements regarding biospecimen collection, emphasizing anonymity, removing potentially confusing or offensive items, and restricting redundant questions.

**Conclusions:** This cognitive interview study offered an empirical pathway for incorporating direct community feedback into the structural and linguistic layout of the PECAN survey. It identified issues related to wording, interpretation, cultural relevance, and respondent comfort informing revisions designed to improve the clarity, comprehensibility, and cultural appropriateness of survey items. While subsequent large-scale psychometric validation is required to establish statistical reliability, this early-phase qualitative process demonstrates the value of community-engaged cognitive interviewing as a formative step in the development of survey instruments intended for diverse populations. Future administration of the revised survey in larger and more diverse samples remains necessary to evaluate its reliability, validity, and broader applicability.

## Background

Addressing health disparities and promoting health equity requires an understanding of the biological and environmental determinants of health. Precision health aims to reduce disparities in chronic diseases like lupus by developing proactive and personalized solutions to health problems. This emerging field integrates inter-individual variability in genetic, behavioral, psychosocial, and environmental determinants of health (1). The collection and sharing of individuals’ unique genomic, physical, chemical, biological and social environmental information is essential for developing effective personalized therapies for everyone. This has been recognized as a national priority, as reflected in federal initiatives such as the All of Us precision health initiative, which aims to build a diverse health database to advance personalized medicine and improve health outcomes for all populations (2, 3). Despite the promise of precision health, significant challenges remain in ensuring that racial and ethnic minority groups, who are often underrepresented in biomedical research, can fully benefit from these advances. Historical mistrust, lack of awareness, and systemic barriers have contributed to the reduced participation of many minority communities to participate in research (4–6). This is especially critical in the context of precision health, where diverse data is essential to develop therapies that are truly inclusive and effective (7). Engaging minority communities in research requires building trust, enhancing awareness, and ensuring culturally sensitive study designs, particularly through community-based participatory approaches, addressing informational needs, and providing direct benefits (8).

One way to ensure that community voices are reflected in research is through the use of cognitive interviewing during the development of survey instruments. Cognitive interviewing is a qualitative method used to explore how respondents interpret, understand, and answer survey questions, helping researchers identify potential issues with wording, cultural relevance, or clarity. This formal technique evaluates whether questionnaire items are understandable and useful for drawing valid conclusions, offering a cost-effective way to improve survey instruments and provide evidence for the content validity of items (9, 10). This method has been successfully employed in public health research to ensure that surveys are accessible and valid across diverse populations.

Given the lack of validated surveys specifically designed to assess beliefs and attitudes about precision health, the Precision rEsearCh pArticipatioN (PECAN) study opted to use cognitive interviews in the development of its survey instrument. The PECAN study is a mixed methods pilot that aims to understand factors influencing participation in precision health research among diverse communities in South Carolina. While the survey is designed for broad community distribution, it oversamples specific target groups impacted by health disparities, including African Americans and individuals with lupus. Since precision health is a relatively new concept, cognitive interviews were essential to ensure that the survey questions would effectively capture participants’ perceptions, beliefs, and concerns regarding research participation in this emerging field.

In this brief report, we describe the cognitive interview process used in the PECAN study to guide the development of a culturally sensitive and scientifically sound survey instrument. We outline how cognitive interviews were conducted, the insights gained, and how this approach facilitated the creation of a more accessible tool for data collection. By incorporating feedback from a diverse sample of participants, the cognitive interview process played a crucial role in ensuring that the final survey instrument would be understandable, culturally relevant, and meaningful for the communities it aims to engage.

## Participants and Methods

### Study Design

The research team developed a survey to identify socioeconomic and cultural factors that influence perceptions about precision health medicine research participation. The survey was specifically designed to assess: 1) familiarity with terminology, 2) perceptions of factors associated with personal health, 3) knowledge, values, and beliefs about genetics research, 4) values considered important when deciding to participate in a research study, and 5) importance of various factors in a policy on precision health medicine research. It asked several questions regarding knowledge, values, and beliefs about precision health research, preferences for how biospecimens and data should be handled, and concerns about participating in precision health research. The survey included a brief description of the study and its purpose, definitions of various precision health-related terms, a demographics questionnaire section, and questions about knowledge, values, experiences, the importance of various factors to be considered when deciding to participate in research, and the importance of various factors to be considered in a policy. Demographic questions included self-reported race, Hispanic/Latino ethnicity, age, gender identity, nativity, educational attainment, current employment, household income, residential status, marital status, and health insurance status. Since different individuals may prefer different terms when describing their race and/or gender, individuals were asked to describe how they prefer to self-report. Since precision health research is a new field and no standard or validated tool exists to measure these concepts, we adapted from previously collected interview questions in diverse groups and community-based settings (11, 12) to explore concerns, values and beliefs related to research (i.e., participation, biospecimen and data storage). Since the team was considering including a question about political affiliation, the participants were asked whether political affiliation could influence participation in precision health research. Racial discrimination was measured using the validated Experiences of Discrimination (EOD) (13) and Everyday Discrimination Scale (EDS) (14) measures. Social Support was assessed using the validated Medical Outcomes Study (MOS) Social Support Survey (15).

A guided group discussion, using a cognitive interview approach, was conducted with community members to improve the clarity and comprehensibility of this research survey. This method allowed participants to verbalize their thoughts while reviewing survey questions, offering real-time feedback on potential issues with the survey instrument. The *“How To” Guide to Cognitive Interviewing* by Gordon B. Willis (9) was used to guide the design of this cognitive interview study. Although concurrent verbal probing using scripted probes was the main technique used, spontaneous probes were also allowed during the interview.

### Participants

We reached out to community members of the Sea Island Families Project (SIFP) Citizen Advisory Committee (CAC) to request volunteers to participate in this study. The SIFP-CAC an academic-community partnership formed between interdisciplinary research teams from the Medical University of South Carolina (MUSC) and Gullah African Americans residing in rural South Carolina that has been supporting community-engaged research projects for over 25 years (16). Four community members, consisting of two men (60 – 70 years old) and two women (30 – 40 and 50 – 60 years old), self-identified as African American, agreed to participate in the study. All participants are actively engaged in efforts to promote and improve access to care among local populations, ensuring their insights were relevant to the objectives of refining the survey instrument. The participants acceptance of the invitation and coming to the scheduled meeting in a café is considered their consent to participate in the cognitive interview. Prior to the interview, the participants were offered a pastry and drink of their choice and following the interview they received $40 USD as compensation for their time, effort, and travel expenses. The Institutional Review Board (IRB) at the Medical University of South Carolina (MUSC) determined that this project is a quality improvement study, and therefore not subject to IRB review or approval.

### Procedure

The cognitive interviews were conducted in a group setting by four interviewers (STK, MAC, DLK, PSR) and lasted about 90 minutes. The draft questionnaire was administered using concurrent cognitive probing, in which the participants were allowed to read each question, and interviewers used guides with a set of scripted verbal probes for assessing any aspect of the questions that the participants found confusing, difficult to answer, or unclear. The probes included: Is the question/statement clear? If so, what makes it clear? If not, what makes it unclear? How would you ask this question to make it clearer? How comfortable would you feel about responding to this statement? What would make you feel comfortable about responding to this statement? What would make you feel uncomfortable about responding to this statement? How would you say these words to make them clearer? Which words would you use? How hard would it be to respond to this statement? What would make it easier to respond to this statement? The discussion was facilitated by a trained moderator (DLK), who also used spontaneous probes to follow-up and prompt participants to elaborate on their feedback. Specific attention was given to identifying potential barriers to comprehension or misinterpretation of survey items.

### Data Collection and Analysis

Four researchers (STK, MAC, DLK, PSR) independently took detailed notes during the cognitive interview session. Each investigator summarized their notes and quotes in a template, which were assembled and summarized by one investigator (STK), enabling the identification of common themes, problems, and key findings. Each researcher initially reviewed their notes to identify recurring issues and areas of concern. The team then met to compare findings, collaboratively identify common themes (e.g. “difficulty understanding,” “hard to answer,” and “ambiguous wording”), and resolve discrepancies through consensus discussion among team members. When differing interpretations emerged, they were addressed through iterative conversation until the team reached a shared understanding. This pragmatic, consensus-based approach allowed for flexible refinement of the survey based on participant feedback. Based on the review of participant feedback, revisions were made to the questionnaire (e.g., wording change, format change, item order change, item removal) to enhance its clarity and relevance to the target population. The resulting survey was pretested among three team members (LAU, DLK, PSR) to ensure that the REDCap platform operated as planned and there were no technical issues.

## Results

### Clarity and Simplification of Language

Participants consistently emphasized the need for clearer, more concise language throughout the survey. For instance, questions involving personal actions, such as “I am willing to undergo genetic testing,” were perceived as too direct and personal, leading to a recommendation to depersonalize the language. Revisions included changing “insurance status” to “insurance coverage” to ensure respondents understood the terms, while ambiguous statements such as “developing drugs that not all can afford” were clarified for better comprehension. Instead of the two original questions with open ended answers (1) “Where do you get your media information?” and (2) “Where do you get your news information?”, the participants recommended clarifying the question by merging into one and offering specific options (TV, Internet, Social apps, Newspaper, Radio, Other) instead of open answer. The majority of participants expressed a preference for simplified phrasing to avoid confusion or misinterpretation of the survey items. The resulting survey is included as Supplementary Material and a selection of questionnaire items that were modified or omitted are highlighted in Table 1.

**Table 1.** Selected questionnaire items with associated interview probe, participant response, and revised survey item.

| Original Questionnaire Item | Cognitive Interview Probe(s) | Participant Response | Issues Identified | Revised Item |
| --- | --- | --- | --- | --- |
| Definitions | How do the definitions make you feel? Which words would you use to make the Definitions clearer? | "Why are you telling me these definitions?"; "Are you going to ask me for specimens? It's scary." | Fear of being asked to provide biological samples for this study | Here are some definitions that may help with terms used in the following survey questions. Disclaimer: We will not be asking you for specimen samples and only want to ascertain your understanding of these definitions. |
| How much do the following items affect your health? | Is this question clear? How would you ask this question to make it clearer? | Noted that 'impact' felt more relevant, and that ranking would be challenging | Preference for "impact" over affect | How important are the following items to your health? |
| I am willing to participate in precision health research | How comfortable would you feel about responding to this statement? | "I am willing to participate.", makes me feel targeted. It seems that these questions are being sneaked in to make me agree to participate and share my info for future research" | Participant felt targeted by the question, concern for coercion and desire for assurance about anonymity and confidentiality | I would be willing to participate in precision health research |
| I have strong spiritual or religious beliefs | What would make it easier to answer this question? | "For what?" | Too vague and does not directly connect to participation in research activities | Omitted |
| Where do you get your media information?<br>Where do you get your news information? | How would you ask this question to make it clearer? | Merge and offer specific options (TV, radio, internet/social media, newspaper). | Lacks clarity | Where do you get your media/news information?<br>(Check all that apply) |

### Context and Reassurance

A common concern was the need for contextual information, especially when introducing potentially sensitive terms like “biological sample” and “biospecimen” under Definitions, at the beginning of the survey. Participants noted fear and apprehension regarding these terms, questioning why these definitions were being shown, and whether they would be asked to provide biological samples for this study. To address this, participants suggested adding a reassuring statement at the beginning of the survey, clarifying that no specimens would be required for this study. Additionally, participants strongly recommended incorporating reminders about the anonymity of responses, particularly in questions related to genetic testing and health insurance, to alleviate concerns about potential impacts on insurability or healthcare access.

### Cultural and Community Representation

Participants highlighted the importance of community representation within the survey. Specifically, the phrasing “researchers should be from the community” was considered too vague and not reflective of respondents’ own communities. Feedback led to the revision of this statement to “researchers should be from *your* community,” reinforcing the need for inclusivity and cultural sensitivity.

The question about having strong spiritual or religious beliefs was found to be confusing, and there was a discussion as to whether clarify, move, or omit. The team decided to omit. Similarly, when asked whether political affiliation could influence participation in precision health research, three of the four participants said that they would be put off by this question, which was not included in the final survey.

### Question Structuring and Merging

Several participants identified redundancy in the survey, particularly between related questions. They recommended merging similar items to improve flow and reduce the cognitive load on respondents. For example, participants proposed combining questions related to the affordability of drugs and the effects of study results on health insurance into a single item. Additionally, questions considered redundant were omitted to streamline the survey. Spiritual beliefs were reassessed, and the specific question ‘I have strong spiritual or religious beliefs’ was ultimately omitted to ensure a more logical progression of topics. When asked about including questions to ask about religious and political views, most participants felt these would be considered offensive. The remaining questions specifically focused on beliefs related to participation in research or biospecimen donation, which is particularly relevant for addressing participant concerns and enhancing engagement in these areas.

### Comprehensive Scope of Health Factors

Participants suggested expanding the scope of the health-related questions to account for a broader range of factors. Specifically, they recommended adding “occupation/job/profession” as a response option to the question addressing determinants of health. This change was suggested to better capture the diverse factors influencing participants’ health, beyond the more commonly addressed variables such as insurance coverage and income.

## Discussion

The cognitive study conducted for the PECAN survey illuminated several critical areas needing improvement, highlighting the importance of refining research instruments through direct feedback from community members. Participants encountered challenges with the original survey’s complex wording and culturally irrelevant content, which led to recommendations for significant revisions to enhance clarity and respondent comfort. Specifically, the study revealed that questions involving personal actions and intricate phrasing, such as those related to genetic testing, were seen as overly direct and potentially invasive. Simplifying the language and depersonalizing these questions were essential steps in making the survey more accessible and less intimidating for respondents.

Additionally, concerns about the implications of terms like “biological sample” and “biospecimen” were prominent among participants, with many fearing these terms implied a request for specimens that could deter their participation. This is consistent with the historical reluctance of the public to give blood for research (17), especially among many African Americans (18). Nevertheless, these concerns do not seem to apply to less invasive biospecimens such as saliva or buccal swabs (17–19). To address these concerns, the cognitive study performed herein recommended incorporating reassuring statements and contextual information at the beginning of the survey, clarifying that no specimens would be requested and emphasizing the anonymity of responses. Such measures are crucial in alleviating participant anxieties and fostering a more comfortable survey-taking experience. Although the PECAN study does not involve biospecimen collection, these questions were intentionally included given the reliance on biosamples in precision health research. Understanding community perceptions of biospecimen use is essential for preparing ethically grounded and culturally responsive research practices in future studies. Participants’ feedback highlights the importance of transparent communication regarding the purpose, handling, and implications of biosample collection, which should be further explored in subsequent phases of this work.

The feedback also underscored the need for greater cultural and community representation within the survey. Participants felt that the original phrasing, such as “researchers should be from the community,” lacked specificity and did not fully acknowledge their own community contexts. Revising this to “researchers should be from *your* community” helped to enhance inclusivity and build trust, ensuring that the survey was more relevant and respectful of the participants’ cultural backgrounds. This aligns with the work done by Lemke *et al* (20), which noted that communities need reassurance that the proposed research would not cause harm, either in the immediate or long-term future, through the researchers’ demonstration of understanding and respect for the community’s perspectives on potential group harms (20). The inclusion of community members as part of the research team is one strategy that can be particularly effective in fostering this understanding, as it promotes bi-directional learning and communication essential for identifying and avoiding potential group harms (21).

Furthermore, the study identified issues with the survey’s structure, including redundancies and cognitive overload. Merging related questions and reorganizing content improved the survey’s flow and reduced the burden on respondents. For example, combining questions about drug affordability and health insurance effects streamlined the instrument, making it more efficient and easier to navigate. Participants also recommended expanding the scope of health-related questions to include factors such as occupation/job/profession, which ensures a more comprehensive range of determinants affecting health is captured. These revisions are especially critical in light of varying levels of health literacy, as advances in precision medicine have outpaced our understanding of patient perceptions across its three domains—genetics, behavioral, and environmental determinants of health (11). Individuals with lower health literacy may struggle to engage with more complex topics, while those with higher literacy may provide deeper insights. By refining and broadening the survey, we can ask more nuanced questions without overwhelming respondents, ensuring that it remains accessible yet capable of collecting meaningful data across different literacy levels, ethnic and racial groups, and determinants of health.

This study has several limitations. Cognitive interviewing is a useful means for testing survey and improving questions, but does not validate questions in a statistical or other formal psychometric way (9). The cognitive interview sample was small and consisted of four community members from a single longstanding community advisory group. While this community-engaged approach provided valuable insights into survey clarity, comprehension, and cultural relevance, this sample size precludes formal psychometric validation, and the findings may not be representative of all populations targeted by the PECAN study or generalizable to broader populations beyond this setting. Although revisions were informed by detailed qualitative feedback from participants, subsequent iterative cognitive re-testing with a separate sample would have strengthened validation of the final instrument’s clarity, comprehensibility, and acceptability. Despite this limitation, the cognitive interview process proved to be a critical, formative, and informative optimization tool for enhancing the quality of the survey. Additionally, while some items were merged or removed to reduce redundancy and respondent burden, we recognize this may limit the ability to assess internal consistency of certain constructs. As this was an early-phase survey refinement step, we plan to evaluate the statistical reliability and validity of the revised instrument during future survey administration to a larger and more diverse cohort. In future phases of the PECAN study, we will consider exploring the use of digital technologies to enhance survey accessibility and engagement. Technology-enabled strategies-such as adaptive surveys, multimedia support, or interactive mobile platforms have the potential to reduce cognitive overload, personalize content delivery, and improve comprehension across varying levels of health literacy. This consideration aligns with prior findings that web-based tools can facilitate participation while introducing both opportunities and challenges in health research engagement (22).

## Conclusion

Overall, this cognitive studyprovided valuable, formative insights that informed refinement of the PECAN survey instrument. Participant feedback led to revisions designed to improve the clarity, comprehensibility, cultural relevance, and organization of survey items. These revisions support the use of community-engaged cognitive interviewing as a valuable approach for developing surveys intended for diverse populations and complex health topics. However, because iterative cognitive re-testing was not performed on a secondary sample, ambiguities may remain, and further empirical administration to a larger, diverse cohort is required to formally establish the statistical reliability, psychometric validity, and long-term utility of the instrument. Ultimately, this process highlights the crucial role of direct participant feedback in developing research tools that are inclusive and culturally accessible.

## Abbreviations

*PECAN*: Precision rEsearCh pArticipatioN

## Supplementary Information

Supplementary Material 1 [PDF document with final REDCap Survey]

## Acknowledgements

The authors would like to thank the volunteers who took the time to participate in the cognitive interview.

## Authors’ contributions

RW wrote the original manuscript draft. BJW and CGA contributed to methodology. LHM, MLG, STK, MAC contributed to investigation. LAU, DLK, and PSR contributed to supervision. PSR contributed to project administration. MAC, CGA, DLK, BJW, and PSR contributed to the conceptualization of this study and funding acquisition. All authors reviewed, edited and agreed on the final version of the manuscript.

## Data availability

The data generated in the current study are available in the supplementary material, and available from the corresponding author upon reasonable request.

## Declarations

### Ethics approval and consent to participate

The Institutional Review Board at the Medical University of South Carolina determined that this project is not considered research under federal regulations, but a quality improvement study, and therefore not subject to IRB review or approval. Since this study only involved a cognitive interview, the United States Department of Health and Human Services regulations for the protection of human subjects (45 CFR part 46) do not apply to such quality improvement activities, and there is no requirement under these regulations for such activities to undergo review by an IRB, or for these activities to be conducted with patient informed consent. The participants acceptance of the invitation and coming to the scheduled meeting in person is considered their consent to participate in the cognitive interview.

### Consent for publication

Not applicable

